# Breast cancer in pregnant young women: clinicohistological profile, risk of death and pregnancy outcomes

**DOI:** 10.1101/2022.09.29.22280276

**Authors:** Ramírez-Torres Nicolás, Rivas-Ruíz Rodolfo, Reyes-López Alfonso, Ureña-Wong Kingston

## Abstract

**Objective:** To describe a set of tumor characteristics, prognosis and course of pregnancy in patients diagnosed with pregnancy-associated breast cancer (PABC).

**Methods:** Retrospective cohort study of PABC young women. The histological profile, survival and pregnancy outcomes were assessed. Nonparametric tests, Fisher’s exact test, Kaplan-Meier method, Cox regression and multivariate logistic regression were used for statistical analyses.

**Results:** We assessed 16 PABC patients. All women self-palpated a breast mass, the women ≤ 35 years of age were diagnosed with unfavorable characteristics: advanced stage (88.8%), positive clinically lymph nodes (100%), high grade (55.5%), ER-negative (77.8%) and high-risk Nottingham prognostic index (66.7%).

Seven deaths were observed with a median follow-up for overall survival (OS) of 64.5 months (range: 15-90). The 5-year OS rates were worse for patients with pathological lymph nodes > 4 (25%; *p* = 0.001) and with ER-negative disease (50%; *p* = 0.646).

In our multivariate analysis, the nodal involvement was the only predictor associated to a worse OS (hazard ratio = 1.4, 90% confidence interval [CI]: 1.14 to 1.8). The following risk factors could influence in the risk of a preterm birth: mother’s older age, gestational age at diagnosis and the chemotherapy during pregnancy, but their adjusted ORs of .61 (90% CI: 0.34 to 1), .80 (90% CI: 0.66 to 0.9) and .01 (90% CI: 0.00 to 0.9), respectively did not support statistically such an effect. Most cases of cases (77.7%) exposed to chemotherapy during pregnancy got a live term birth.

**Conclusion:** Our findings described a more aggressive histological profile for youngest pregnant women coupled the delayed diagnosis might explain the high-risk of death. Simultaneous management of breast cancer and pregnancy was feasible.

## Background

Breast cancer is one of the most common malignancy diagnosed among pregnant women, obstetricians are seeing a higher number of cases with pregnancy-associated breast cancer (PABC) ^1-3^ due to the upward trend of the first childbearing after the third decade of life combined with the increased incidence of the breast cancer worldwide ^2-11^.

This entity is uncommon ^2-14^, it is estimated at 1/3,000 to 1/10,000 pregnancies, instead for the developed countries the incidence rises varying from 15 to 44 per 100,000 pregnancies ^2-5^. The diagnosis of breast cancer during pregnancy represents a true challenge, justifies a multidisciplinary physician team to plan better therapeutic strategies for both the mother and the unborn child ^7-11^. An extensive research has been carried out improving the knowledge about the tumor characteristics, oncological management and obstetrical-fetal cares in PABC patients ^2-26^. However, the prognosis is remains subject of debate due to conflicting outcomes in PABC studies ^2-6, 11-26^.

Physiological changes during pregnancy increase mammary nodularity and engorgement, which might mask signs and symptoms of early breast cancer ^2, 9, 12-14, 19^, making more difficult the clinical and radiologic diagnosis. We should avoid the misdiagnosis of any breast mass since might contribute in a delayed diagnosis of tumors.

This research described tumor characteristics, survival and pregnancy and neonatal outcomes, the information might help to any physician to begin a prompt intervention and refer to PABC patients to specialist physicians of breast and pregnancy.

## Patients and methods

### Characteristics of the study population

This study is a descriptive cohort involving women in whom cancer was diagnosed during pregnancy with an observational-based between March 1992 and June 2010. All cases were under the care of a multidisciplinary physician team (composed of oncologists, obstetricians, anesthesiologists, internists and neonatologists) who work at Ginecology and Obstetrics Hospital No. 3, National Medical Center “La Raza”, belonging Mexican Institute of Social Security, it is a tertiary reference center for pregnant women and breast cancer.

Pregnant women who were ≤ 40 years of age were included with histological diagnosis of unilateral breast cancer in clinical stage I to III. Imaging studies for diagnosis and staging were carried out in all patients according to standard guidelines to ensure fetal health. The clinical stage was based on TNM (tumor, node, metastasis) classification for solid cancers of the AJCC (6th. edition) ^27^. The TNM stage previous to 2002 was restaged. We used the traditional definition for PABC, it is the breast cancer diagnosed during pregnancy or within one year after the delivery ^7-15^.

### Clinicohistological factors

Epidemiological, clinicohistological and immunohistochemistry (IHC) data were gathered from medical records which were described in detail previously ^26^. When T, N and grade were combined, risk groups were established according to the Nottingham prognostic index (NPI) criteria ^21, 28^. Subsequently, we arbitrarily divided into two groups the NPI: low/intermediate-risk NPI (< 5.4 score) and high-risk NPI (> 5.4 score).

A retrospective IHC analysis was performed for the estrogen receptor (ER), progesterone receptor (PgR) and human epidermal growth factor receptor 2 (HER2) without Ki67. We used these IHC surrogate indicators and the grade to classify approximately the tumors in five molecular subtypes as other studies have done ^29, 30^: luminal A (LA), luminal B/HER2-negative (LB/HER2-), luminal B/HER2-positive (LB/HER+), HER2-positive (HER2+) nonluminal and triple negative (TN).

### Oncological management

The information of the standardized oncological management (chemotherapy, definitive surgery and radiotherapy), course of pregnancy and perinatal outcomes were described previously ^26^. Almost all patients received the same drugs (5-fluorouracil, epirubicin and cyclophosphamide both in the neoadjuvant chemotherapy (NC) and adjuvant chemotherapy (AC) by back then time.

### Statistical analysis

The main objective was to describe clinicohistological characteristics with their prognostic influence on survival in pregnant women with breast cancer. The second objective was to assess the disease free survival (DFS) and overall survival (OS) curves.

We compared the histological profile between two age groups (≤ 35 years of age and > 35 years of age) using the Fisher’s exact test. Medians and IQR (interquartile range: 25th-75th) were assessed using nonparametric tests for age, tumor size and NPI. The Kaplan-Meier method assessed the survival function and the groups were compared using the log-rank test ^29-31^. The hazard function was also assessed. All alive patients without events were censored at the time of last follow-up.

The associations between prognostic factors and hazard of death were assessed using Cox proportional hazards regression for OS. Multivariate models estimated the adjusted hazard ratio (HR), regression coefficient (β) and confidence intervals (CI) ^31^. Person-time at risk was counted from date of breast cancer diagnosis until date of death or censoring. We hypotheised: more aggressive clinicohistological factors are more frequent in PABC younger patients.

The birth (term or preterm) was associated to risk factors for pregnancy using multivariate logistic regression, the odds ratio (OR) was estimated with the null hypothesis of no relationship. A significance level of 10% was used in Cox regression and logistic regression. STATA version 14 (College Station, TX) was used for all statistical analyses. The study was assessed by the institutional review board.

## Results

### Clinical characteristics

Sixteen PABC patients were assessed. The epidemiological and histological data are presented in Table 1. The median age at presentation was 35 years (IQR: 34-36). Of 16 patients, nine cases were ≤ 35 years of age, three (18.8%) cases had family history of breast cancer and at least three-quarters had history of a previous pregnancy.

**Table 1.** Clinical characteristics of pregnant woman with breast cancer

| Characteristic | n = 16 (%) |
| --- | --- |
| Age (years) |  |
| Median | 35 |
| Interquartile range | (34 - 36) |
| Age by group |  |
| < 30 years | 2 (12.4) |
| 31-35 years | 7 (43.8) |
| > 36 years | 7 (43.8) |
| Tumor size (cm) |  |
| Median | 6 |
| Interquartile range | ( 5 - 7.5) |
| Tumor size (T) |  |
| T2 | 4 (25.0) |
| T3 | 8 (50.0) |
| T4 | 4 (25.0) |
| Axillary lymph nodes (N) |  |
| N0 | 1 ( 6.2) |
| N1 | 9 (56.2) |
| N2 | 6 (37.5) |
| Clinical stage |  |
| IIA | 1 ( 6.2) |
| IIB-IIIA | 11 (68.7) |
| IIIB | 4 (25.0) |
| Histology |  |
| Ductal carcinoma | 12 (75.0) |
| Lobular carcinoma | 1 ( 6.2) |
| Medullary carcinoma | 1 ( 6.2) |
| Other carcinoma | 2 (12.4) |
| Histological grade |  |
| Grade 1 | 1 ( 6.2) |
| Grade 2 | 8 (50.0) |
| Grade 3 | 7 (43.8) |
| Lymphovascular invasion |  |
| Absent | 10 (62.5) |
| Present | 6 (37.5) |
| History of breast cancer |  |
| No | 13 (81.2) |
| Yes | 3 (18.8) |

The median of the primary tumor at diagnosis was 6 cm (IQR: 5-7.5). Most the cases showed a tumor larger than 5 cm (75%), axillary clinical nodal involvement (93.7%), infiltrating ductal carcinoma (75%) and locally advanced stages at diagnosis (IIB onward: 93.7%). Table 1. No inflammatory breast cancer was observed. Only one pathological complete response after NC was obtained.

### Clinical characteristics by age group

Pregnant women ≤ 35 years of age were diagnosed with high percentages of more aggressive prognostic factors: clinical stage III (88.8%), positive clinically lymph nodes (100%) and grade 3 tumor (55.5%). Regarding IHC markers, tumors classified as ER/PgR-negative predominated in 77.8% and 100%, respectively and one third was HER2+ tumor. But the distribution of histological profile did not differ significantly between both age groups (all *p* > 0.05). Table 2.

**Table 2.** Baseline characteristics by age group

| Characteristic | Status | Age,<br>< 35 years | Age,<br>> 35 years | *P* * |
| --- | --- | --- | --- | --- |
|  |  | n = 9 (%) | n = 7 (%) |  |
| Clinical stage | IIA-B | 1 (11.1) | 2 (28.6) | 0.550 |
|  | IIIA-B | 8 (88.8) | 5 (71.4) |  |
| Lymph nodes | N0 | 0 ( 0.0) | 1 (14.2) | 0.192 |
|  | N1-N2 | 9 (100.) | 6 (85.4) |  |
| Tumor grade | G1-G2 | 4 (44.4) | 5 (71.4) | 0.358 |
|  | G3 | 5 (55.5) | 2 (28.6) |  |
| ER | Positive | 2 (22.2) | 5 (71.4) | 0.126 |
|  | Negative | 7 (77.8) | 2 (28.6) |  |
| PgR | Positive | 0 ( 0.0) | 4 (57.1) | **0.019** |
|  | Negative | 9 (100.) | 3 (42.9) |  |
| HER2 | Negative | 6 (66.7) | 5 (71.4) | 1.000 |
|  | Positive | 3 (33.3) | 2 (28.6) |  |
| NPI | < 5.4 | 3 (33.3) | 4 (57.1) | 0.615 |
|  | > 5.4 | 6 (66.7) | 3 (42.9) |  |

ER: estrogen receptor; PgR: progesterone receptor; HER2: human epidermal growth factor receptor 2; NPI: Nottingham prognostic index. \* Fisher's exact test.

### Survival of the study population

For all patients, the median follow-up for DFS was 47.5 months (range: 0-81) and for OS was 64.5 months (range: 15-90). The survival curves are shown in figures 1A to 1F; using the log-rank test, the number of pathological lymph nodes (pNs) influenced negatively at DFS (5-year: 29% vs. 100%; *p* < 0.001) and OS (5-year: 25% vs. 100%; *p* = 0.001) for pNs > 4 and pNs of 0-3 subgroups, respectively. Figures 1A and 1D.

**Figure 1.**
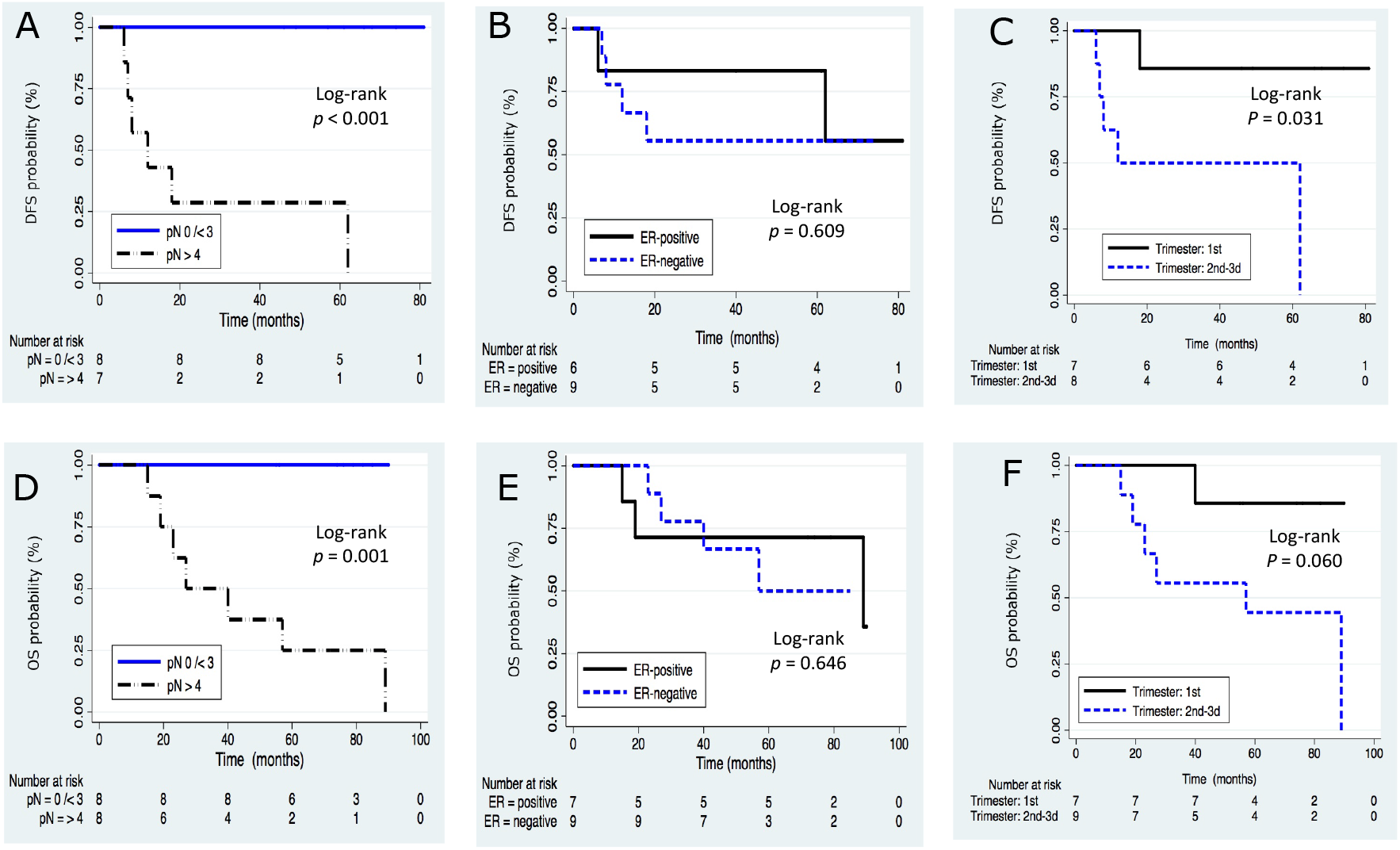
Differences in Kaplan-Meier curves according to pathological lymph nodes (pNs), estrogen receptor (ER) and trimester of pregnancy at diagnosis of breast cancer were observed. A, B, C for disease-free survival (DFS). D, E, F for overall-survival (OS).

Patients with ER-negative tumors showed a trend toward a shorter survival at 5 years than for patients with ER-positive tumors, the DFS was 56% vs. 83% (*p* = 0.609) and OS was 50% vs. 71% (*p* = 0.646), respectively. The differences did not reach statistical significance for both DFS and OS. Figures 1B and 1E.

Patients in whom cancer was diagnosed during the first (1st.) trimester of pregnancy favored significantly a better DFS at 5 years compared with those diagnosed during second (2nd.) and third (3rd.) trimesters (86% vs. 50%, respectively; *p* = 0.031). Although there was an improved OS for 1st. trimester group (86% vs. 44%, respectively; *p* = 0.060), no statistical significant difference was observed. Figures 1C and 1F.

### Risk of death

We assessed the effect of prognostic factors on survival. Table 3. Bivariate analyses ascertained that the lymph node involvement and NPI experienced a 35% and 90% increase in the hazard of death, their HRs were 1.35 (90% CI: 1.13 to 1.6) and 1.9 (90% CI: 1.10 to 3.2), respectively. In multivariate analysis after adjustment for selected pronostic factors, only the lymph node status affected significantly OS, by each additional lymph node in the number of pNs increased a 44% the hazard of death (90% CI: 1.14 to 1.8) compared with lower number of pNs. The other variables were less clear on OS. Table 3.

**Table 3.** Cox regression analysis of covariables affecting OS in pregnant women with breast cancer Bivariate analysis Multivariate analysis

| Covariable | Bivariate analysis |  |  |  | Multivariate analysis |  |  |  |
| --- | --- | --- | --- | --- | --- | --- | --- | --- |
| | $\beta$ | SE | HR (90% CI) | *p* | $\beta$ | SE | HR* (90% CI) | *p* |
| pN (number) | 0.301 | 0.107 | 1.35 (1.13-1.6) | **0.005** | 0.367 | 0.144 | 1.44 (1.14-1.8) | **0.011** |
| Age (years) | -0.245 | 0.131 | 0.78 (0.63-0.9) | 0.062 | -0.224 | 0.137 | 0.79 (0.63-1.0) | 0.102 |
| Tumor (cm) | 0.006 | 0.160 | 1.00 (0.77-1.3) | 0.969 | -0.253 | 0.346 | 0.77 (0.43-1.3) | 0.463 |
| ER-negative | 0.398 | 0.872 | 1.48 (0.35-6.2) | 0.648 | -0.598 | 1.309 | 0.54 (0.06-4.7) | 0.648 |
| NPI (score) | 0.638 | 0.329 | 1.90 (1.10-3.2) | 0.052 |  |  |  |  |
| Trimester<sup>£</sup> | 1.788 | 1.082 | 5.98 (1.01-35.) | 0.099 |  |  |  |  |

OS: overall survival; $\beta$ : regression coefficient; SE: standard error; HR: hazard ratio; CI: confidence interval; pN: pathological lymph node;
ER: estrogen receptor; NPI: Nottingham prognostic index; trimester of pregnancy at diagnosis<sup>£</sup>: 1st. vs. 2nd. -3d.
HR\*: adjusted for pN, age, tumor and ER

### Risk of preterm birth

The following risk factors could influence at increased risk of preterm births: mother’s older age, gestational age at diagnosis and chemotherapy during pregnancy, but their adjusted ORs of .61 (90% CI: 0.34 to 1.1), .80 (90% CI: 0.66 to 0.9) and .01 (90% CI: 0.00 to 0.9), respectively did not support statistically such an effect, maybe due to a statistical underpowered sample. Figure 2. The most cases of cases (77%) exposed to chemotherapy during pregnancy had a live term birth.

**Figure 2.**
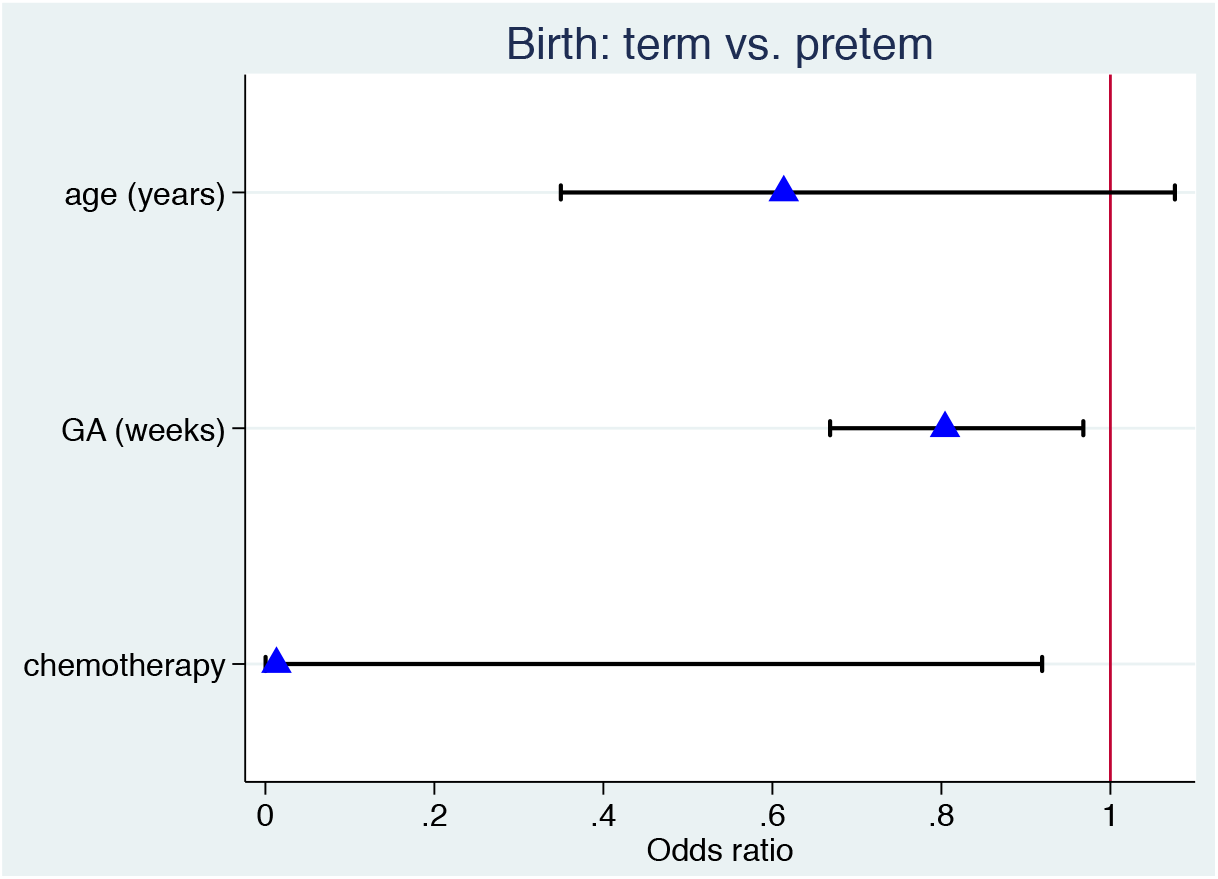
Forest plot of logistic regression for birth adjusted for mother’s age, gestational age (GA) at diagnosis and chemotherapy (during postpartum vs. pregnancy) in pregnant women with breast cancer

### Follow-up

We observed a rate of 939 months-person, during this time frame there were seven deaths among PABC patients with locally advanced stage (IIB to IIIB), of this group four were grade 3 tumors. The most common molecular subtypes that developed distant disease were HER2+ (four out of five) and TN tumors (three out of six).

Of nine PABC patients exposed to chemotherapy during pregnancy, a third had distant recurrences after chemotherapy completion, only one case had progression and death during the treatment with pregnancy interruption. The main sites of metastasis were: lung (five cases), bones (four cases) and brain (one case), only two cases had local recurrence. All patients (seven) with distant disease have died.

## Discussion

The presence of any neoplasm in pregnant woman faces clinical dilemmas and challenges in the cancer management and pregnancy cares ^7-11, 13, 32-34^. Establishing questions such as: when should it began chemotherapy, what drugs can be used during pregnancy, what are the best obstetrical-fetal strategies favoring the health of the dyad mother-child ^7-11, 17, 32-34^.

In this study, a third (37.5%) of women who were ≤ 30 years of age delayed their first pregnancy, of this group the half were nulliparous women, most PABC patients (81.2%) had chilbearing history. None pregnant patient reported an abortion induced or a pregnancy interrupted. The family history of breast cancer was not associated with PABC since only three cases had this history ^26^.

The prognostic factors are often taken in account to establish the prognosis. The TNM stage is one of the most important parameters influencing on survival, breast cancer during pregnancy is often diagnosed at locally advanced stages (IIb onward) accounting for 39% to 90% ^11-14, 19-21, 23-26^, which partly explains the poor prognosis.

In this cohort, all women self-detected a palpable breast mass, almost all women were diagnosed at locally advanced stage (93.7%: IIB to IIIB) which may explain a poor survival. Patients with larger tumors had increased hazard of death, but the unadjusted HR reduced its magnitude of 1.0 to 0.77 (90% CI: 0.43 to 1.3) after adjustments in the multivariare model, **so tumor size did not influence in the hazard of death**. Perhaps due to lack of variability, since three-quarter of patients had tumors larger than 5 cms.

The lymph node involvement often occurs in PABC patients in more than two thirds of cases (50% to 93.7%) ^6, 12-26^, this prognostic factor is considered as the strongest predictor in the prognosis. In our cohort, the clinical lymph node involvement of ipsilateral axillary region was a relatively high clinical finding (93.*7*%). The number of axillary pNs was stratified following NC-surgery or surgery alone. Half of cases had pNs < 3, the other half with pNs > 4 exhibited a high percentage of distant recurrences of 87.5% (seven out of eight). The unadjusted HR for nodal status increased of 1.35 to 1.44 after the adjusments in the multivariate model, we were able to confirm a strong effect of the lymph node involvement on HRs estimates, so patients with higher number of pN are more likely to die.

Young age at diagnosis is considered a poor prognostic factor, since has an increased risk of distant recurrence and death compared to middle and older-aged women ^15, 29, 35-38^. Several studies have found a high mortality rate in younger women, particularly in ER-positive and luminal tumors ^16, 25, 29, 35^. PABC patients compared with those non-PABC are more as likely to be diagnosed at younger age, with advanced T/N stages, lymph node involvement, high grade and ER/PgR-negative tumors ^7-15, 25, 26, 32-34^. TN tumors ^6, 16-18, 21, 25, 29, 35-37^ and BRCA1/2 gene mutations ^7, 8, 36^ are far more common in younger women. All these combined parameters establish a high risk of recurrence and death.

The younger group is associated with a worse prognosis due to high expression of unfavorable biohistological features and the most PABC patients are young women. So, pregnant young woman with more aggressive histological profile is recommended chemotherapy after the 1st. trimester to reduce the risk of recurrence and death ^7-11, 26, 32-34^.

In this study, we found an increased mortality rate (56%) in women ≤ 35 years of age compared with those > 35 years of age (29%), perhaps is influenced by the delayed diagnosis of tumor. However, the point estimate remained unchanged in bivariate and multivariate analyses (HR = 0.79, 90% CI: 0.63 to 1.0). Thus, the age did not impact as the main driver of the increased mortality rate.

Other tumor features have been described for PABC women, the predominant histology is the infiltrating ductal carcinoma (accounting for 75% to 93%) ^2-6, 11-26^, medullary breast cancer is infrequent representing of 2.*7*% to 6.*2*% ^6, 7, 23^. Most studies have reported grade 3 tumors ranging from 48% to 84% ^6, 12-18, 20-22, 25^, lymphovascular invasion is identified in 21% to 67.3% of cases ^12-14, 16-18, 20, 21, 26^.

Most studies have found in PABC patients a higher frequency of ER-negative (45% to 62%) and PgR-negative tumors (35% to 59%) ^6, 11-13, 16-19, 23, 24, 39^, both receptors are also commonly negative which are associated with a poor prognosis ^6, 19, 21, 25, 37, 39^.

In this study, although the ER-negative tumor was common (77.8%) in women younger than age 35 years subgroup, but the mortality rate was similar for both ER-negative and ER-positive tumors (44% vs. 43%, respectively). The unadjusted HR for ER-negative reduced its magnitud of 1.48 to 0.54 (90% CI: 0.06 to 4.7) after adjustments in the multivariate model, so that we found null association between the ER status and mortality.

The HER2 overexpression (17% to 65%) ^6, 12, 14-18, 20, 21, 23-26, 39^ and high levels of ki-67 (39% to 100%) ^12, 16-18, 21, 23-25^ are reported with wide variability. Few studies reported the p53 abnormal expression accounting for 48% to 68.3% ^12, 21, 23^, in view of inconsistent data, we should continue evaluating these IHC biomarkers in PABC patients.

The main IHC biomarkers (ER, PgR, HER2, ki67) have been incorporated to investigate the molecular subtype ^16-18, 20, 23-25^. PABC young women often present TN tumors (28% to 50%) ^2, 7, 14, 16-18, 21, 25^. Some Asian ^23, 24^ and European ^20^ studies have reported a higher frequency of luminal B subtype. Thus, the molecular subtypes differ according to the race and geographical region.

In this study we found higher frequency of TN (37.5%) and LA (25%) tumors, but HER2+ tumors (with or without LB tumor profile) showed a higher mortality rate over other subtypes, exhibited three critical points in mortality rate, one maximum at 25 months and two minimums at 40 and 55 months for both subtypes, respectively. Figure 3.

**Figure 3.**
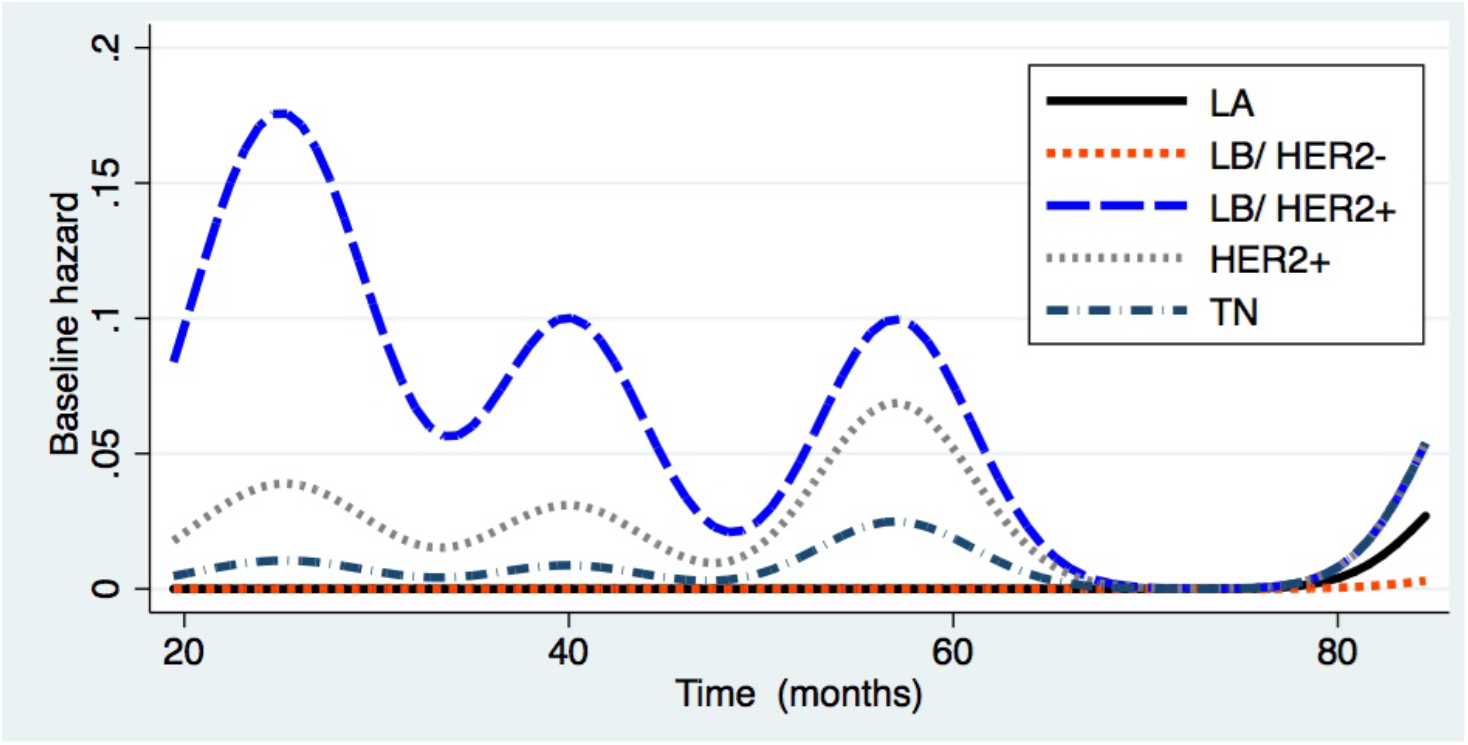
Mortality rate according to molecular subtype on overall survival. LA: Luminal A; LB: Luminal B; HER2: human epidermal receptor factor 2; TN: Triple negative

Multiple case-control studies ^19-25^, population-based cohorts ^2, 3, 5, 6, 37^ as well as meta-analysis studies ^40-42^ have ascertained an increased risk of death if the breast cancer is diagnosed during pregnancy. A wide research suggests that tumors diagnosed in all time frames of the postpartum exhibited differences in the tumor biology and a significant increased risk of death, even the risk is more pronounced for those diagnosed within 12 months postpartum compared with those non-pregnancy-related breast cancer ^2, 4, 6, 37, 40-46^; some consider the postpartum can extend up to 5 or 10 years after the delivery ^6, 37, 40-46^; the physiological events of pregnancy and postpartum are intertwined, but both entities should be separated because there are wide differences both biohistological features and in prognosis ^4, 6, 37, 40-46^. We should also consider whether patients already had a subclinical disease prenatal.

For example in cases-control studies, Rodriguez et al. ^19^ and Liao et al. ^24^ noted that PABC patients had 14% and 23% increased risk of death compared to controls, the HRs were 1.14 (95% CI: 1.0 to 1.29) and 1.23 (95% CI: 0.46 to 3.29), respectively. Azim et al. ^20^ noted an inferior OS as well for pregnant women with HR of 2.6 (95% CI: 1.0 to 6.5).

In a meta-analysis of 45 studies, Shao et al. ^42^ assessed 6,602 cases and 157,657 controls for DFS and OS. PABC patients compared to controls significantly showed increased risk of death with a pooled hazard ratio (pHR) of 1.45 (95% CI, 1.30 to 1.63), the heterogeneity was significant (*I* ^2^ = 64.9; *p* < 0.001). The mortality was significant for those diagnosed at 12 months after the last delivery (pHR = 1.59, 95% CI: 1.3 to 1.82). Instead, the mortality was not significantly different at 70 months after the last delivery (pHR = 1.14, 95% CI: 0.99 to 1.25).

Whereas other studies found similar survival rates in PABC patients compared to the nonpregnant control group ^4, 11, 13-18, 37^ after matching by age and stage. For example the OS at 5 years showed by O’Sullivan et al. ^11^ was 75% and 81%, by Amant et al. ^15^ was 78% and 81% and by Ploquin et al. ^17^ was 83.1% and 85.5%, respectively (all *p* > .05).

Beadle et al. ^13^ noted a similar at 10-year survival for both PABC (64.6%) and non-PABC (64.8%%). Instead, Litton et al. ^14^ found contrarian results, OS was slightly superior in the PABC women than for controls (77% vs. 71%, respectively; *p* = 0.046). It is not clear exactly why they found improved outcomes in the pregnant patients.

Only a subgroup of PABC women are more likely to have a worse prognosis; the age, nodal status and stage are not the only factors implicated to exert a poor prognosis. It is not clear whether hormonal effects and immune changes induced during pregnancy or postpartum might be implicated upon exerting an adverse effect over stroma favoring the proliferation of mammary stem cells ^2, 20, 43, 47-49^ or already existing malignant clones ^43, 47^.

In rodent models, the breast involution process during the postpartum has been considered as a potential mediator of tumor growth ^45-51^, structural changes in the mammary tissue microenvironment including the extracellular matrix remodeling, fibrillar collagen deposition, macrophages infiltration, important proteolysis, increased angiogenesis ^52^, among other events that resemble wound healing, might promote tumor accelerated growth and dissemination ^43-51^ which predicts a poor prognosis in young women ^47^.

It can not be ruled out that mortality is influenced by pregnancy’s own factors ^7, 20^, it is possible that the tumor cells may have a more pronounced adverse effect after a more recent pregnancy at an older age ^6, 43, 44^. Moreover, immune infiltrate of the breast cancer cells may contribute in the tumor biology and prognosis ^20, 43, 46, 51, 53^.

PABC patients share many tumor features occurring in nonpregnant women ^2-6, 11-25^; pregnancy may contribute to a delay in diagnosis and management of breast cancer and the poor prognosis might be explained by the delayed diagnosis of breast cancer ^6-11, 20, 25, 26, 43^ or because the tumors tend to show a more aggressive biology. In our study, the breast cancer diagnosed during 2nd. and 3rd. trimesters was associated with almost sixfold higher hazard of death (unadjusted HR = 5.9, 90% CI: 1.01 to 35) than those diagnosed during 1st. trimester, but no significant difference on OS was observed.

The NPI also predictes risk groups in PABC women or in the postpartum ^21, 38^. However, Lambertini at el. ^54^ described that the Adjuvant! Online model is a reliable tool in predicting OS at 10 years, whereas the performance of NPI is sub-optimal.

In our study, the NPI median value (6.1) was classified as poor prognosis. Patients with NPI higher score significantly had a 90% increased hazard of death (unadjusted HR = 1.9, 90% CI: 1.1 to 3.2) compared with NPI lower score. Thus, the individual analysis of classical prognostic factors is not recommend to establish the ultimate prognosis ^21, 38, 53^.

Weaknesses of our study should be mentioned, possible misclassification of the tumor measurement during pregnancy, data were gathered over a long period and the major limitation was the small number of cases. Therefore, there may be limitation to establish firm conclusions. Instead the uniformity highlights in oncological management for PABC patients who received the same epirubicin-based chemotherapy during the 2nd. and/or 3rd. trimesters or in the postpartum, in addition all were followed at same hospital that ensured the analysis of complete outcomes of the breast cancer and pregnancy ^26^.

Any palpable mass in the breast or axillary region that persists for longer than two weeks during pregnancy should be examined, the biopsy helps us to establish the histological diagnosis. Breast cancer during pregnancy often presents as a painless mass or thickening in the breast. The management of cancer during the development of high-risk pregnancy should be individualized taking into account patient health status, TNM stage and gestational age at diagnosis ^7-11, 32-34^, since the application of conventional therapies is not possible at all times ^7, 15, 20, 32-34^.

The direct interaction among young age, pregnancy and breast cancer can be simultaneously managed. Regarding surgery, the 2nd. trimester is the safest period to perform a surgery, although this may be performed irrespective of gestational age ^7-11, 26, 32-34^. In our study, we initially underwent a breast-conserving surgery without axillary lymphadenectomy in 14 patients and modified radical mastectomy (MRM) in two cases during pregnancy. After the delivery, the major surgery (MRM) underwent in the postpartum in eight cases.

The use of chemotherapy can be associated with increased risk of obstetrical and fetal complications, such as preterm labor, premature rupture of membranes and the intrauterine growth restriction. The transient myelosuppressive effect of chemotherapy affects both the mother and the child, which should be treated based on the standard recommendations. Anthracyclines and alkylating agents can be safely used after 1st. trimester and hormonotherapy after the completion of chemotherapy if indicated ^7-11, 32-34^.

In our institution, the chemotherapy was applied in 56% of cases (seven as NC and two as AC) during the 2nd. and/or 3rd. trimesters with supportive medication (ondansetron and corticosteroids) to reduce side effects. Serious side events or deaths in mothers or neonates exposed to chemotherapy were not observed ^26^. The radiotherapy was postponed until after delivery only in nine cases.

Regarding pregnancy, for all cohort the median gestational age at the delivery was 37.5 weeks (range: 10-40 weeks), 81.2% of cases ended in live birth and three cases ended in miscarriage. Patients who received chemotherapy during pregnancy, at least three-quarter of cases (77.7%) had a live term birth and two had a live preterm birth. Delivery after 37 weeks is recommended, avoiding premature delivery whenever possible and vaginal delivery should be favored over cesarean section ^7-11, 15, 32-34^. In our study was not so, the elective cesarean section underwent in ten cases and the vaginal delivery in three cases.

Regarding child, the perinatal cares always were avaliable, nine newborns had birth body weight above 2,500 g and four a birth body weight below 2,500 g. The Apgar score > 8 at 1 minute was assessed in ten cases and Apgar score < 8 in three cases; almost all neonates were healthy except for two cases with respiratory failure, one died by extreme underweight and prematurity (interruption of pregnancy at week 31 due to disease progression), the other child was alive after the physician cares ^26^. Newborns did not show apparent congenital malformations and their neurophysiological development have been normal in the first three years after the delivery reported by their parents ^26^.

Regarding birth, thanks multidisclipinary pyshician team, our institution obtained a high percentage of term births, reducing the admission likely to intensive cares for preterm newborns, perhaps because all patients with breast cancer associated with high-risk pregnancy at diagnosis had closer cares to reduce the perinatal morbidity. Although, PABC itself is a risk factor for a preterm birth.

The epidemiological and histological characteristics and TNM stage are similar to other local study ^25^. Our study confirmed that women ≤ 35 years of age subgroup with PABC tended to exhibited a more aggressive immuno-histological profile such as advanced T/N stage, high grade, ER/PgR-negative, high-risk NPI and TN tumor as well as the delayed diagnosis might almost fully explain the poor prognosis. Our findings were concordant with other studies that document more aggressive tumor characteristics and ultimate prognosis in PABC patients ^2-6, 11-25, 32-34, 37, 45^.

## Conclusion

Simultaneous management of breast cancer and pregnancy was feasible. Our experience institutional supports that the breast cancer can be relative safety treated during pregnancy and closer obstetrical monitoring must be carried out until its term to get optimal outcomes: healthy mothers with term births without complications. Therefore, we should continue planning better specialized strategies to maximize health outcomes for both.

## Data Availability

All data produced in the present study are available upon reasonable request to the authors

## Acknowledgements

The authors thank all patients who shared their information, social workers Elvira and Sara for greatly helping in getting the medical records and librarian Jorge Pérez for his technical support in the physician journals. We also thank Clinical Research Training Center of Mexico city for the support in methodology of this research.

